# Quantifying the Quality of Corrective Actions in Medical Safety Incident Reports Using a Hybrid Rule-Based and Large-Language-Model Classification System: A Cross-Sectional Pilot Feasibility Analysis of 11,507 Japanese National Reports (2010, Interim)

**DOI:** 10.64898/2026.08.04.26359750

**Authors:** Hitoshi Sugawara

**Affiliations:** Department of Family & General Medicine, Tokyo-Kita Medical Center, Tokyo, Japan; Division of General Medicine, Department of Comprehensive Medicine 1, Jichi Medical University Saitama Medical Center, Saitama, Japan

**Keywords:** patient safety, incident reporting, corrective action, medical errors, prevention & control, safety management, quality improvement, health informatics, natural language processing, large language models, text classification

## Abstract

**Background:** Whether corrective actions documented in medical safety incident reports rely on individual vigilance (“Safety-I”) or on structural, system-level intervention (“Safety-II”) has not been quantitatively evaluated on a national scale in Japan. We developed an automated classification pipeline to assign corrective-action free-text to a 7-level maturity scale (L0–L6) and computed two summary indices: the Safety Measure Quality Profile (SMQP), the full L0–L6 distribution, and the System-based Safety Measure Rate (SSMR), the proportion of non-L0 records classified L3–L6.

**Methods:** We analyzed all 11,507 corrective-action free-text entries from the 2010 release of Japan’s national medical accident and near-miss reporting database (Japan Council for Quality Health Care, JCQHC), comprising 8,804 near-miss (Hiyari-Hatto) and 2,703 accident (Jiko) reports. Records were classified using a five-stage hybrid pipeline: an expert-developed rule dictionary, TF-IDF + k-nearest-neighbor matching, cosine-similarity matching, a two-tier large-language-model (LLM) classifier, and a conservative priority-cascade fallback. SSMR was compared between near-miss and accident reports using a χ^2^ test, Wilson 95% confidence intervals, Cramér’s V, and the risk difference (RD), against pre-specified minimal clinically important difference (MCID) criteria of RD ≥ 2 percentage points and Cramér’s V ≥ 0.10.

**Results:** Every record received a definitive L0–L6 label (0% unresolved). Overall, 16.6% of records were unclassifiable (L0); among the 9,599 classifiable (non-L0) records, individual-vigilance actions (L1) predominated (54.6% of all records), and only 11.82% (95% CI, 11.19–12.49%) met the SSMR criterion (L3–L6). SSMR was higher for accident reports than for near-miss reports (18.12% [95% CI, 16.69–19.64%] vs. 9.46% [95% CI, 8.80–10.17%]; RD = 8.66 percentage points; Cramér’s V = 0.120; χ^2^(1) = 136.97, *p*<0.001), exceeding both pre-specified MCID thresholds.

**Conclusions:** In this interim single-year analysis, the large majority of documented corrective actions in Japanese medical safety reports remained individual-vigilance-based rather than system-based, with accident reports showing a substantively, rather than merely statistically, higher proportion of system-based actions than near-miss reports. These findings support the feasibility of large-scale automated assessment of corrective-action quality and provide the rationale for the planned 16-year longitudinal analysis.

**Status note:** This report presents an interim, single-year (2010) result generated during the rule-dictionary development and validation phase of a planned longitudinal study spanning 2010–2025 (16 years, N = 178,375; UMIN000071271). This pilot report corresponds to Study 2 (SMQP/SSMR: corrective-action quality assessment) of the planned longitudinal study and is intended to evaluate methodological feasibility before application to the complete 2010–2025 corpus. Findings reported here should not be interpreted as the study’s primary longitudinal outcome and are subject to revision once the classification dictionary is frozen and applied to the full corpus.

## Introduction

Patient-safety practice has progressively shifted from an individual-vigilance model, in which harm prevention depends on a clinician’s attention or memory, toward a systems-oriented model in which harm is prevented by redesigning the work environment itself (1, 2). National incident-reporting systems such as Japan’s Medical Accident Information Collection Project, operated by the Japan Council for Quality Health Care (JCQHC) (3), accumulate large volumes of free-text corrective-action descriptions, but whether these actions have moved beyond individual-level exhortation toward structural, system-level intervention has not been quantified on a national scale. Previous studies using JCQHC data, including those by Akiyama and colleagues (4) and by Onishi and colleagues (5), have focused on specific aspects of incident reports rather than the quality of corrective actions themselves. Consequently, whether incident-report systems predominantly generate individual-vigilance measures or system-based interventions remains unclear. Automated classification of incident-report text using large language models has recently been demonstrated at scale (6-8); the present pipeline extends this general approach from event classification to grading the maturity of the corrective actions.

We developed a 7-level classification scheme (L0, unclassifiable; L1, individual vigilance; L2, education/communication; L3, procedure/documentation; L4, standardized verification; L5, environment/equipment; L6, forcing function), broadly aligned with the NIOSH Hierarchy of Hazard Controls (9), together with two derived indices: the Safety Measure Quality Profile (SMQP, the full L0–L6 distribution) and the System-based Safety Measure Rate (SSMR, the proportion of non-L0 records classified L3–L6). This report presents a pilot feasibility analysis of the 2010 reporting year (N = 11,507), conducted during an ongoing dictionary-development and validation process for a planned 16-year (2010–2025, N = 178,375) longitudinal study. The findings are intended to assess methodological feasibility and provide a baseline reference before a future frozen dictionary is applied to the full corpus.

## Methods

### Reporting guideline

This study was reported in accordance with the Strengthening the Reporting of Observational Studies in Epidemiology (STROBE) statement for cross-sectional studies(10). A completed STROBE checklist, cross-referenced to the relevant sections of this manuscript, is provided as the Supplementary Table S1.

### Study design and data source

This is a pilot feasibility study based on a cross-sectional analysis of a single reporting year (2010), conducted as an interim step within a planned longitudinal study (UMIN000071271; ethics approval:

Tokyo Kita Medical Center Clinical Research Ethics Review Committee, approval no. 558, approved 2 July 2026). We used the publicly available 2010 release of the JCQHC Medical Accident Information Collection Project, which comprises two report types: near-miss (“Hiyari-Hatto”) and accident (“Jiko”) reports. A participant flow diagram was constructed according to STROBE recommendations (Figure S1). Of 11,507 reports available in the 2010 JCQHC release, all contained non-empty corrective-action fields and were included in the analysis (N = 11,507; Hiyari-Hatto, n = 8,804; Jiko, n = 2,703); no records were excluded, and all records ultimately received a final classification label. Because this is a secondary analysis of de-identified, publicly released aggregate report data with no patient-, reporter-, or institution-identifying information, the requirement for informed consent was waived.

No formal sample size calculation was performed because all eligible reports available in the 2010 JCQHC release were included; this pilot analysis was therefore population-based for the selected year rather than sample-based.

### Classification pipeline

Corrective-action free text was normalized (Unicode NFKC normalization and full-width/half-width harmonization) and processed through a five-stage hybrid classification pipeline. The stages were applied sequentially, with each stage acting only on records unresolved by the preceding stage. (1) an expert-developed rule dictionary (regular-expression pattern matching, rule-dictionary MD5 hash: 4226564766ffabbe831004cbed54c761 used to ensure reproducibility of dictionary versions); (2) TF-IDF character n-gram (2–4) features with k-nearest-neighbor classification (k = 7, cosine metric, accepted only at predictive probability ≥ 0.55); (3) cosine-similarity matching against previously classified records (accepted at similarity ≥ 0.85); (4) a two-tier LLM classifier (a lower-cost model for routine cases, escalating low-confidence or longer cases to a higher-capacity model, temperature = 0); and (5) a conservative priority-cascade fallback (evaluated in the order L6→L5→L4→L3→L2→L1, defaulting to L0 only when no signal matched any level). Every record therefore received a definitive L0–L6 label; no record remained unclassified. Full pattern-matching rules, including the pre-specified requirement that isolated mentions of “double-check” map to L1 (individual vigilance) unless accompanied by explicit structural-establishment language, are not publicly available during the ongoing dictionary-development and validation phase but are planned for release following completion of the full study.

### Outcome definitions

The Safety Measure Quality Profile (SMQP) is the full distribution of records across L0–L6, operationally defined in Table 1. The System-based Safety Measure Rate (SSMR) is defined as:

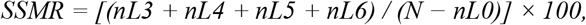

where nLx denotes the number of records assigned to level Lx.

**Table 1.**
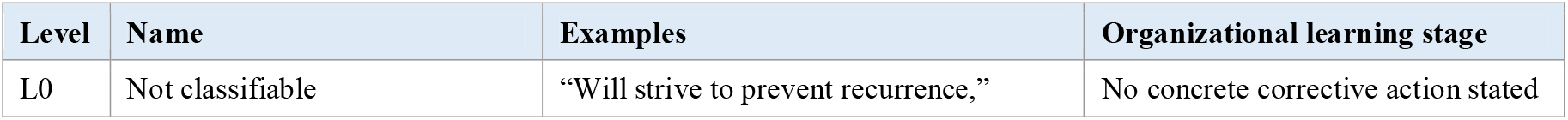
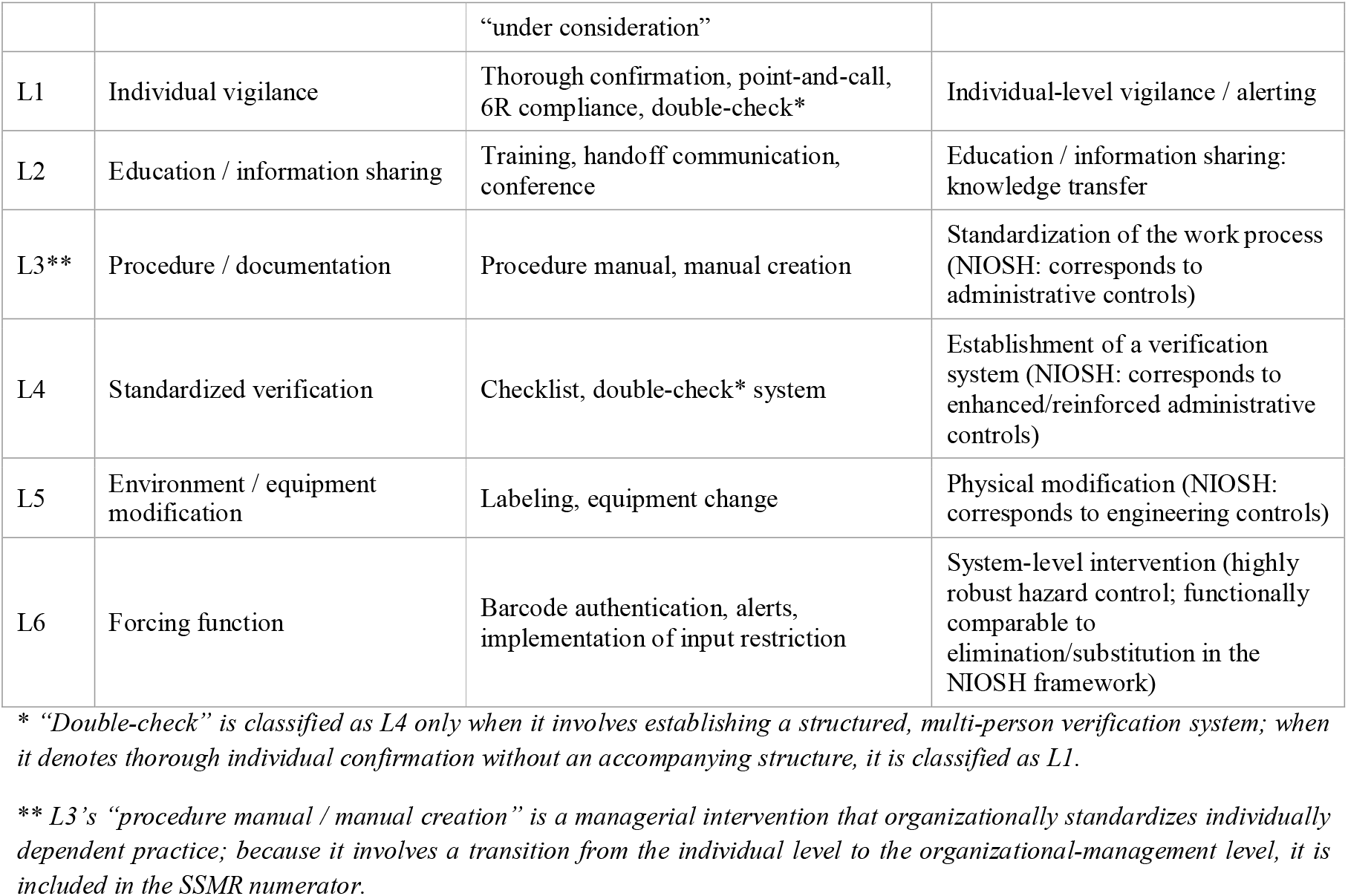
Operational definitions of the L0–L6 classification levels.

i.e., the proportion of classifiable (non-L0) records assigned to a system-based level (L3–L6). L0 (unclassifiable; no concrete corrective action stated) is excluded from the SSMR denominator by design, since it reflects absence of an evaluable corrective action rather than a maturity level. L0 prevalence is reported separately as a component of the SMQP.

### Bias mitigation

Several measures were implemented to reduce classification bias. First, deterministic rules were applied before machine-learning or LLM-based methods whenever possible, so that the most transparent and auditable component of the pipeline took precedence. Second, all thresholds used in the KNN, cosine-similarity, and LLM classification stages (Methods, Classification pipeline) were predefined before any statistical comparison between report types was performed. Third, statistical analyses were conducted only after the classification process was finalized to avoid outcome-driven refinement of the classification algorithm.

### Statistical analysis

SMQP and SSMR (with Wilson 95% confidence intervals) were computed overall and separately for near-miss and accident reports. The two report types were compared using a χ^2^ test on the 2×2 contingency table of report type by SSMR status among non-L0 records, together with Cramér’s V, the risk difference (RD), and the relative risk (RR). MCID thresholds were specified a priori: an absolute risk difference of 2 percentage points was selected as the minimum operationally meaningful effect — approximately six times the standard error of the overall SSMR estimate in this dataset (SE ≈ 0.33 percentage points, N = 9,599) — whereas Cramér’s V ≥ 0.10 corresponded to the conventional threshold for a small effect size (11). No missing data were present in the analyzed variables, because only reports containing a non-empty corrective-action field were included; consequently, no imputation procedures were required. Sensitivity analyses (e.g., alternative treatment of L0 records, rule-only versus full-pipeline classification) are planned for the full-corpus study but were not performed in the present single-year pilot analysis. As this is a single-year, cross-sectional pilot, no trend or change-point analysis was performed; longitudinal trend estimation is deferred to the planned 16-year analysis. All analyses were performed in Python 3.12 using pandas 2.2.2, NumPy 2.0.2, and SciPy. A global random seed of 42 was used throughout the analytical pipeline to enhance reproducibility.

## Results

All 11,507 eligible corrective-action records from the 2010 JCQHC release were successfully classified. No record remained unresolved after the five-stage pipeline (Table 2). The rule dictionary alone resolved 74.5% of records (n = 8,570, including 89 records classified through an additional vocabulary layer introduced during manual quality-control review); the remaining records were resolved by k-nearest-neighbor matching (22.7%, n = 2,606), the LLM classifier (2.9%, n = 330), or cosine-similarity matching (<0.1%, n = 1). No record required the fallback classifier.

**Table 2.** Cohort characteristics, JCQHC 2010 release.

| Characteristic | Near-miss<br>(Hiyari-Hatto)<br>$n = 8,804$ | Accident<br>(Jiko)<br>$n = 2,703$ | Overall<br>$N = 11,507$ |
| --- | --- | --- | --- |
| Corrective-action text length, median (IQR), characters | 33 (12–62) | 74 (40–124) | 40 (17–76) |
| Records classified by rule dictionary, $n$ (%) | — | — | 8,570 (74.5%) |
| Records classified by KNN / cosine / LLM, $n$ (%) | — | — | 2,937 (25.5%) |
| Records unresolved (fallback required), $n$ (%) | — | — | 0 (0.0%) |

Corrective-action descriptions were substantially longer in accident reports than in near-miss reports (median 74 vs. 33 characters).

The overall SMQP is shown in Figure 1. L1 (individual vigilance) was the single most common level, accounting for 54.6% of all records (n = 6,279); L0 (unclassifiable) accounted for 16.6% (n = 1,908); L2 (education/communication) accounted for 19.0% (n = 2,185). System-based levels (L3–L6) together accounted for only 9.86% of all records (n = 1,135): L3, 2.03%; L4, 3.11%; L5, 4.23%; L6, 0.49%. Overall SSMR — the proportion of classifiable (non-L0) records reaching a system-based level — was 11.82% (95% CI, 11.19–12.49%; 1,135/9,599).

**Figure 1.**
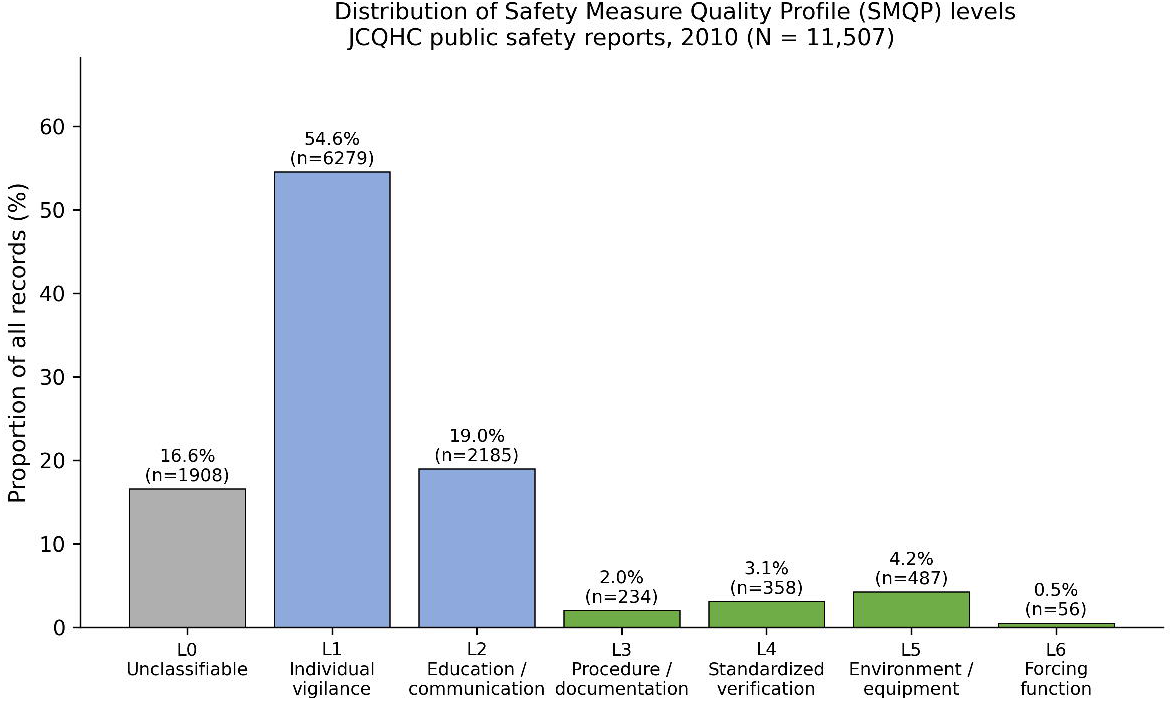
Distribution of Safety Measure Quality Profile (SMQP) levels across all 2010 records (N = 11,507). Bars are shaded by broad category: unclassifiable (grey), individual/educational actions (light blue, L1–L2), and system-based actions (green, L3–L6).

SMQP differed materially between report types (Figure 2, Table 3). SSMR was higher for accident reports than for near-miss reports (18.12% [95% CI, 16.69–19.64%] vs. 9.46% [95% CI, 8.80–10.17%]; RD = 8.66 percentage points; χ^2^(1) = 136.97, *p*<0.001; Cramér’s V = 0.120), meeting both pre-specified MCID criteria (RD ≥ 2 pp and Cramér’s V ≥ 0.10). Accident reports were approximately twice as likely to contain a system-based corrective action as near-miss reports (RR = 1.92, 95% CI, 1.72–2.14). Accident reports also had a markedly lower proportion of unclassifiable (L0) entries than near-miss reports (3.0% vs. 20.8%).

**Table 3.** SMQP distribution by report type, JCQHC 2010 release.

| Level | Near-miss, $n$ (%) $n = 8,804$ | Accident, $n$ (%) $n = 2,703$ | Overall, $n$ (%) $N = 11,507$ |
| --- | --- | --- | --- |
| L0 | 1,827 (20.75) | 81 (3.00) | 1,908 (16.58) |
| L1 | 4,969 (56.44) | 1,310 (48.46) | 6,279 (54.57) |
| L2 | 1,348 (15.31) | 837 (30.97) | 2,185 (18.99) |
| L3 | 123 (1.40) | 111 (4.11) | 234 (2.03) |
| L4 | 241 (2.74) | 117 (4.33) | 358 (3.11) |
| L5 | 263 (2.99) | 224 (8.29) | 487 (4.23) |
| L6 | 33 (0.37) | 23 (0.85) | 56 (0.49) |
| SSMR, % (95% CI) | 9.46 (8.80–10.17) | 18.12 (16.69–19.64) | 11.82 (11.19–12.49) |

**Figure 2.**
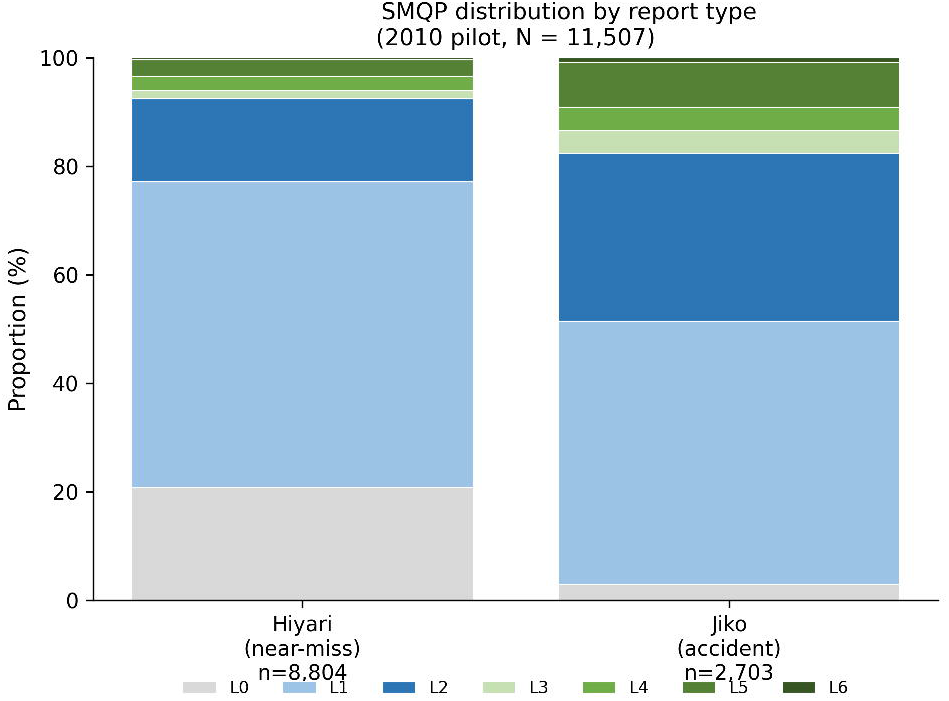
SMQP distribution by report type. Near-miss (Hiyari-Hatto, n = 8,804) and accident (Jiko, n = 2,703) reports, each stacked to 100%.

## Discussion

In this interim, single-year analysis of 11,507 Japanese national medical safety reports, only 11.82% of classifiable corrective actions reached a system-based level (L3–L6); more than half (54.6%) relied on individual vigilance alone (L1). Accident reports showed a substantively higher SSMR than near-miss reports (18.12% vs. 9.46%; RR = 1.92), a difference that exceeded both the pre-specified statistical significance threshold and the minimal clinically important difference thresholds.

This pattern is consistent with concerns raised in both the Safety-II literature and patient-safety reporting research that incident-reporting systems can generate large volumes of documented “lessons learned” without a commensurate shift toward structural redesign of the care environment (2, 12, 13) . The higher SSMR observed for accident reports than for near-miss reports may reflect that more severe events prompt more thorough institutional review and more resource-intensive corrective action, though this cross-sectional, single-year analysis cannot establish causality or directionality, and residual confounding by event type, clinical area, or reporting culture cannot be excluded.

The classification pipeline resolved every record without requiring the fallback classifier, and the rule dictionary alone, the most transparent and auditable component of the pipeline, accounted for three-quarters of all classifications. This suggests the deterministic component of the pipeline is already reasonably mature for a single reporting year, though its performance on the full 16-year, 178,375-record corpus, which spans a wider range of vocabulary and reporting conventions, remains to be established. Importantly, the absence of fallback assignments should not be interpreted as redundancy of the fallback mechanism. Rather, it suggests that the preceding rule-based, similarity-based, and LLM-assisted stages achieved complete classification coverage within this pilot dataset. Nevertheless, the fallback classifier remains part of the production pipeline as a safety-net mechanism designed to guarantee complete classification coverage when all preceding stages fail to produce a classification, and to accommodate vocabulary drift and reporting heterogeneity anticipated across the full 2010– 2025 corpus.

Limitations are noted in the interest of transparency and to guide the planned longitudinal analysis. First, this is a cross-sectional snapshot of a single year (2010); no inference about temporal trend is intended or supported by these data. Second, the rule dictionary underlying this analysis had not yet been frozen at the time of this report; it remains in an active development and validation phase spanning representative time points across the full 2010–2025 corpus. Consequently, the SSMR estimates presented here may change once the dictionary is finalized and the full corpus is processed under a single frozen rule set. Because dictionary development remains ongoing, the rule set and associated source code have not yet been publicly released; full external reproducibility will therefore require confirmation after publication of the final frozen version. Third, a data-integrity review conducted for this report identified that 89 of 11,507 records (0.8%), resolved by an additive vocabulary layer introduced during manual quality-control review, had been recorded under a provenance label that did not map cleanly to the intended “rule” pipeline stage; these records have been reclassified to the rule-dictionary stage in the reporting above, and the underlying stage-mapping logic will be corrected before the frozen production run. Fourth, formal inter-rater reliability statistics (e.g., Cohen’s κ against independent human annotation) for this specific 2010 run are not yet available and will be reported alongside the full-corpus analysis. A target Cohen’s κ of ≥ 0.75 was specified a priori in the parent study protocol. External validation using an independent institutional incident-report dataset is planned as part of the parent study and was beyond the scope of the present pilot analysis. Fifth, because reporting requirements, safety culture, and incident-classification systems differ across countries, direct extrapolation of the present estimates to non-Japanese reporting systems should be undertaken cautiously; nevertheless, the methodological framework may be transferable to other national incident-reporting databases. Finally, because no record reached the fallback stage in the 2010 dataset, the practical utility of that component could not be evaluated empirically; its contribution should therefore be reassessed after application to the full 2010–2025 corpus, where greater vocabulary diversity and reporting heterogeneity are expected.

In conclusion, this pilot feasibility analysis demonstrates the practicality of automated, reproducible classification of corrective-action quality at national scale and provides a 2010 reference point indicating that system-based corrective action remained uncommon in that year. The planned frozen-dictionary analysis of the full 2010–2025 corpus (N = 178,375) will determine whether this pattern has changed over the intervening 16 years. Subsequent phases of the parent study will further evaluate external validity through application of the same classification framework to institutional incident-report data and explore its potential use as a benchmarking tool for assessing the maturity of safety-improvement practices across healthcare organizations.

## Data Availability

The source data analysed in this study are publicly available from the Japan Council for Quality Health Care (JCQHC) Medical Accident Information Collection Project. Aggregate results are contained within the manuscript and supplementary materials. The classification dictionary, analysis code, and derived datasets are not publicly available at the time of this pilot report because the dictionary remains under active development and validation, but are planned for release following completion of the full study.

## Data and Code Availability

Source incident-report data are publicly available from the Japan Council for Quality Health Care (JCQHC) Medical Accident Information Collection Project. The classification pipeline, rule dictionary, and analysis code are not publicly available at the time of this pilot report because the dictionary remains under active development and validation. Public release is planned following completion of the full study and finalization of the production dictionary. The pilot-phase rule dictionary used for this analysis is identified by MD5 hash 4226564766ffabbe831004cbed54c761 (2010 records only; run timestamp: 2026-08-02T03:10:09 UTC).

## Funding

No specific funding was received for this work. As no external funding was received, no funder had any role in study design, data collection, analysis, manuscript preparation, or the decision to submit the work for publication.

## Conflicts of Interest

The author declares no conflict of interest relevant to this work.

## Author Contributions

H.S.: Conceptualization, Methodology, Software, Formal Analysis, Investigation, Writing – Original Draft, Writing – Review & Editing.

## Notes

### Author Declarations

Tokyo-Kita Medical Center Clinical Research Ethics Review Committee of Tokyo-Kita Medical Center gave ethical approval for this work (approval no. 558; approved on 2 July 2026). The requirement for informed consent was waived because this study used publicly available de-identified data and involved no identifiable patient information.

