## Supplemental Table S1 for "Quantifying the Quality of Corrective Actions in Medical Safety Incident Reports Using a Hybrid Rule-Based and Large-Language-Model Classification System: A Cross-Sectional Pilot Feasibility Analysis of 11,507 Japanese National Reports (2010, Interim)"

**Supplementary Materials**

**Table S1.** STROBE checklist for cross-sectional studies

| **Item** | **STROBE recommendation** | **Reported in** |
| --- | --- | --- |
| 1a | Indicate the study's design in the title or abstract | Title |
| 1b | Informative and balanced summary in the abstract | Abstract |
| 2 | Background / rationale | Introduction, ¶1 |
| 3 | Objectives | Introduction, ¶2 |
| 4 | Present key elements of study design early in the paper | Methods, Study design and data source |
| 5 | Setting, locations, and relevant dates | Methods, Study design and data source |
| 6 | Eligibility criteria, sources and methods of selection of participants | Methods, Study design and data source; Figure S1 |
| 7 | Clearly define all outcomes, exposures, predictors, potential confounders, and effect modifiers | Methods, Outcome definitions; Table 1 |
| 8 | Data sources / measurement | Methods, Classification pipeline |
| 9 | Efforts to address potential sources of bias | Methods, Bias mitigation |
| 10 | Study size | Methods, Study design and data source (final paragraph) |
| 11 | How quantitative variables were handled | Methods, Outcome definitions; Statistical analysis |
| 12a-b | Statistical methods, including subgroup/interaction analyses | Methods, Statistical analysis |
| 12c | How missing data were addressed | Methods, Statistical analysis (none present) |
| 12e | Sensitivity analyses | Methods, Statistical analysis |
| 13a-c | Numbers of individuals at each stage of the study | Figure S1 (flow diagram) |
| 14a-c | Descriptive data (characteristics, exposures) | Table 2; Results, ¶1 |
| 15 | Outcome data / summary measures | Table 3; Results, ¶2 |
| 16a-c | Main results, precision, and (if relevant) relative and absolute risk | Results, ¶3 (RD, RR, 95% CI) |
| 17 | Other analyses performed | Not applicable in this pilot (see Statistical analysis / Limitations) |
| 18 | Key results, summarized with reference to study objectives | Discussion, ¶1 |
| 19 | Limitations, discussing bias and imprecision | Discussion, Limitations ¶ |
| 20 | Cautious overall interpretation | Discussion, ¶2–3; Conclusion |
| 21 | Generalisability of the study results | Discussion, Limitations ¶ (fifth point) |
| 22 | Source of funding and role of funders | Funding |
