## Supplementary Figure S1 for "Quantifying the Quality of Corrective Actions in Medical Safety Incident Reports Using a Hybrid Rule-Based and Large-Language-Model Classification System: A Cross-Sectional Pilot Feasibility Analysis of 11,507 Japanese National Reports (2010, Interim)"

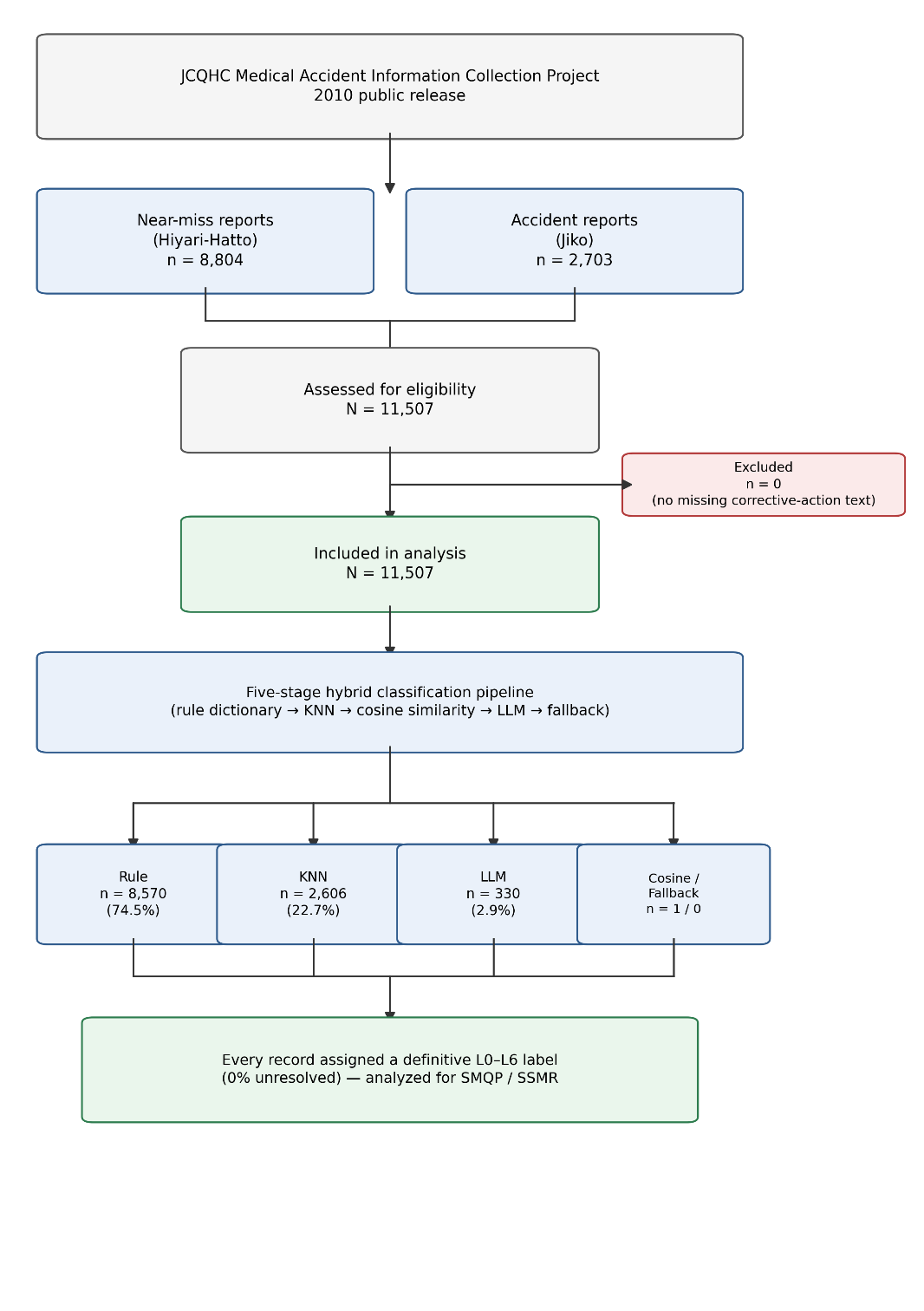

**Figure S1.** Participant flow diagram (STROBE Item 13). All 11,507 reports in the 2010 JCQHC release had non-empty corrective-action text and were included; no records were excluded. Every included record received a definitive L0–L6 label via the five-stage classification pipeline.
